# Pharmacogenomic and drug interaction risk associations with hospital length of stay among Medicare Advantage members with COVID-19

**DOI:** 10.1101/2021.05.06.21256769

**Authors:** Kristine Ashcraft, Chad Moretz, Chantelle Schenning, Susan Rojahn, Kae Vines Tanudtanud, Gwyn Omar Magoncia, Justine Reyes, Bernardo Marquez, Yinglong Guo, Elif Tokar Erdemir, Taryn O. Hall

**Affiliations:** Invitae Corporation, 1400 16 St, San Francisco, CA 94103; OptumLabs at UnitedHealth Group, 5995 Opus Parkway, Minnetonka, MN 55343

**Keywords:** COVID-19, pharmacogenomics, medication management, hospitalization, precision medicine, length of stay, healthcare administration, healthcare costs

## Abstract

**Importance:** COVID-19 has severely impacted older populations and strained healthcare resources, with many patients requiring long periods of hospitalization. Reducing the hospital length of stay (LOS) reduces patient and hospital burden. Given that adverse drug reactions are known to prolong LOS, unmanaged pharmacogenomic risk and drug interactions among COVID-19 patients may be a risk factor for longer hospital stays.

**Objective:** The objective of this study was to determine if pharmacogenomic and drug interaction risks were associated with longer lengths of stay among high-risk patients hospitalized with COVID-19.

**Design:** Retrospective cohort study of medical and pharmacy claims

**Setting:** Administrative database from a large U.S. health insurance company

**Participants:** Medicare Advantage members with a first COVID-19 hospitalization between January 2020 and June 2020, who did not die during the stay.

**Exposures:** (1) Pharmacogenetic interaction probability (PIP) of ≤25% (low), 26%–50% (moderate), or >50% (high), which indicate the likelihood that one or more clinically actionable gene-drug or gene-drug-drug interactions would be identified with testing; (2) drug-drug interaction (DDI) severity of minimal, minor, moderate, major, or contraindicated, which indicate the severity of an interaction between two or more active medications.

**Main Outcomes and Measures:** The primary outcome was hospital length of stay. Results were stratified by hierarchical condition categories (HCC) counts and chronic conditions.

**Results:** A total of 6,025 patients hospitalized with COVID-19 were included in the study. Patients with moderate or high PIP were hospitalized for 9% (CI: 4%–15%; p < 0.001) and 16% longer (CI: 8%–24%; p < 0.001), respectively, compared to those with low PIP, whereas RAF score was not associated with LOS. High PIP was significantly associated with 12%–22% longer lengths of stay compared to low PIP in patients with hypertension, hyperlipidemia, diabetes, or COPD. Finally, among patients with 2 or 3 HCCs, a 10% longer length of stay was observed among patients with moderate or more severe DDI compared to minimal or minor DDI.

**Conclusions and Relevance:** Proactively mitigating pharmacogenomic risk has the potential to reduce length of stay in patients hospitalized with COVID-19 especially those with COPD, diabetes, hyperlipidemia, and hypertension.

**Key Points:** *Question:* What is the impact of unmanaged pharmacogenomic risk among patients hospitalized with COVID-19?

*Findings:* Among 6,025 patients hospitalized with COVID-19, those with greater unmanaged pharmacogenomic risk for adverse drug reactions had longer hospital stays than those with lower risk, both within the entire cohort and within groups matched by number and type of chronic conditions.

*Meaning:* Preemptive pharmacogenomic testing may shorten hospital stay by reducing adverse drug reactions among seriously ill patients and more broadly improve patient risk classification, care utilization predictions, and health system performance.

## Introduction

Hospitalizations represent nearly one-third of all U.S. healthcare expenditures.^1^ Patient length of stay (LOS) is a longstanding measure of hospital efficiency as longer LOS is associated with greater resource utilization and higher costs.^2–5^ Among many factors associated with longer LOS are adverse drug reactions (ADRs), which are preventable with appropriate medication management.^6–8^ Patient risk factors for ADR are inappropriate polypharmacy as well as harboring genetic variants that affect medication response. Pharmacogenomics-guided medication management could therefore reduce both ADRs and LOS. Studies suggest that >99% of individuals harbor at least one variant associated with an atypical drug response and 24% of individuals may currently take a drug to which they may have an atypical response.^9–11^ Given that greater pharmacogenomic variant burden has been associated with longer LOS,^12^ unmanaged pharmacogenomic risk may be an intervenable vulnerability for reducing LOS among high-risk patients.

Population ADR risk management could benefit hospital systems at any time and may benefit hospitals even more so when there is additional population-wide stress on the hospital system, such as the COVID-19 pandemic. The COVID-19 pandemic placed unprecedented demands for hospitals across the globe, and patient demand for hospitalization exceeded hospital resources in several regions.^13–17^ In the US, roughly half of those hospitalized with COVID-19 have been Medicare and Medicare Advantage beneficiaries and the majority of these beneficiaries have been hospitalized for more than 5 days.^18,19^ Medication management interventions may especially help the Medicare population as 54% of patients 65 and older report taking four or more prescription drugs daily.^20^ This increases their risk for ADR-related hospitalization, which is reported to be 4x greater among patients 65 years of age or older compared to younger patients.^21^ Further, polypharmacy is a reported risk factor for developing COVID-19.^22^ Thus, hospitalized COVID-19 patients may be at heightened risk for ADR.

A recent study found that nearly 90% of patients hospitalized with COVID-19 were found to have at least one medication order with pharmacogenomic guidance and nearly one-quarter had orders for 4 or more actionable medications.^23^ However, it is not clear whether pharmacogenomic risk or drug-drug interaction risk affects LOS among COVID-19 patients. To evaluate the impact of pharmacogenomic risk on LOS among high-risk patients, we conducted a retrospective analysis of administrative medical and pharmacy claims among Medicare Advantage members hospitalized with COVID-19. The main objective of this study was to determine if pharmacogenomic or drug-drug interaction (DDI) risk among Medicare beneficiaries hospitalized with COVID-19 was associated with LOS. We further examined this potential association by the number of diagnosed chronic conditions as well as four specific chronic conditions: chronic obstructive pulmonary disease (COPD), diabetes, hyperlipidemia, and hypertension.

## Methods

### Data Source and Study Population

Data were obtained from an administrative database of medical and pharmacy claims from a large U.S. health insurance company. The claims include the International Classification of Diseases, Tenth Revision, Clinical Modification (ICD-10-CM) diagnosis codes, the National Drug Codes (NDC), patient demographic data, and other information. The de-identified data used in this study were granted a waiver of authorization by the institutional review board of the UnitedHealth Group Office of Human Research Affairs.

Claims data for 10,206 Medicare Advantage members admitted to a hospital with COVID-19 between January 1, 2020 and June 30, 2020 were reviewed. COVID-19 diagnoses were identified through SARS-CoV-2 lab test records indicated by the Logical Observation Identifiers Names and Codes (LOINC) organization’s guidance for mapping SARS-CoV-2 and COVID-19-related LOINC terms.^24^ Test information provided via LOINC terms indicated the test type (antibody, RT-PCR, etc.) as well as the test result (detected, not detected, not given/cancelled). Suspected COVID-19 inpatient cases were manually reviewed daily by clinical staff via clinical notes to determine an individual’s COVID-19 status, flagging it as either negative, confirmed, presumed positive, or needing clinical review. All findings except for confirmed status were reviewed daily. Only patients’ first COVID-19 admission was included and evaluated, and patients who died during their hospital stay (n=1,604, or 16% of patients reviewed) were excluded from the study to remove potential confounders. Only patients continuously enrolled in Medicare Advantage Prescription Drug plans for at least 12 months prior to the index COVID-19 admission were eligible for inclusion.

### Independent and Outcome Variables

The primary independent variables were pharmacogenetic interaction probability (PIP) and DDI severity as determined by Invitae’s YouScript clinical decision support tool. PIP is the probability that pharmacogenomic testing of 14 genes (*CYP2C19, CYP2C9, CYP2D6, CYP3A4, CYP3A5, CYP2B6, CYP4F2, DPYD, HLA-B*57:01, IFNL3, SLCO1B1, TPMT, UGT1A1*, and *VKORC1*) will result in the detection of one or more clinically significant pharmacogenomic interactions involving one or more medications on the patient’s current medication list. PIP was calculated based on publicly available phenotype prevalence data in the U.S. population and pharmacogenomic interaction data, which encompasses clinically actionable drug-drug or drug-drug-gene interactions in CPIC Guidelines and FDA labeling. More robust descriptions of YouScript are available in US patents.^25–27^ PIP was categorized into the following: 1) low (PIP ≤ 25%), 2) moderate, (PIP 26%–50%); and 3) high (PIP >50%).

DDI indicates the potential for adverse interactions among 2 or more medications prescribed to a patient, irrespective of the patient’s pharmacogenomic profile. DDI severity was measured on an ordinal scale of increasing severity: minimal, minor, moderate, major, and contraindicated, with moderate or higher being tied to evidence indicating a need to consider changing a drug or dose. Among patients with multiple DDIs in their drug regimen, only the most severe DDI was used in this study.

All medications active or prescribed within 30 days prior to and during the first COVID-19 admission were screened for PIP and DDIs. Other independent variables were age, gender, race, residential location, median income, risk adjustment factor (RAF), HCC, and Special Needs Plan (SNP) type. RAF scores and HCC, which are measures of disease burden used to determine likely need for medical care, were based on the one-year window prior to the patient’s admission month. HCC are simplified diagnosis categories that are constructed using a standardized ICD-10 code mapping determined by the Centers for Medicare and Medicaid Services (CMS). HCC counts indicate the number of chronic conditions and were categorized as 0 or 1, 2 or 3, 4 or 5, or 6 or more, following previously reported CMS categorizations.^28^ HCC, along with demographic data, were used in an algorithm to assign an RAF score, which is a measure of patient complexity.^29^ SNPs are Medicare plan types restricted to individuals with specific diseases or characteristics. SNP types were (1) chronic conditions SNP (C-SNP), for individuals with severe or disabling chronic conditions; (2) dual-eligible SNP (D-SNP), for individuals entitled to both Medicare and state Medicaid; and (3) institutional SNP (I-SNP), for individuals who need or are expected to need services in a long-term care skilled nursing facility or similar high-level care facility for 90 days or longer.^30^ The outcome variable of interest was LOS during the first COVID-19 admission.

## Statistical Analysis

Zero-truncated negative binomial analyses were used to assess the effects of independent variables on LOS. Two sets of analyses were carried out. The first focused on the entire cohort and included the following models: (i) the baseline model that accounted for age, gender, PIP and DDI; and (ii) the regression model that adjusted for the variables identified by least absolute shrinkage and selection operator (LASSO) as covariates with strong association with LOS.^31^ The second set of analyses were subgroup analysis to limit the effects of underlying disease through model fitting and variable selection for each of the following:

1. patients stratified by HCC counts (i.e. number of chronic conditions);
2. patients stratified by each of the following chronic conditions regardless of other comorbidities: (i) COPD, (ii) diabetes, (iii) hyperlipidemia, and (iv) hypertension.

In both sets of analyses, the point and confidence-interval estimate of the rate ratios and the corresponding *P*-values resulting from the fitted models were obtained. Statistical significance was set at *P*<0.05. Additionally, descriptive statistics were applied to profile the study population, including count, percentage, mean, and standard deviation (SD). All analyses were conducted using the statistical computing software R version 3.6.1 (R Foundation for Statistical Computing, Vienna, Austria).^32^

## Results

A cohort of 6,025 individuals met all inclusion criteria, including hospitalization with COVID-19 and 12-month continuous Medicare Advantage coverage (Figure 1). Most patients were female (61%), had a mean age of 77 years (SD, 11 years), were of white (non-Hispanic) ancestry (62%), lived in urban areas (47%), and had an average median household income of $63,027 (SD, $17,435) (Table 1). One-third of patients were enrolled in an I-SNP (34%), and 76% had 2 or more chronic conditions. More than half of the study population had hyperlipidemia (58%) or diabetes (52%).

**Figure 1.**
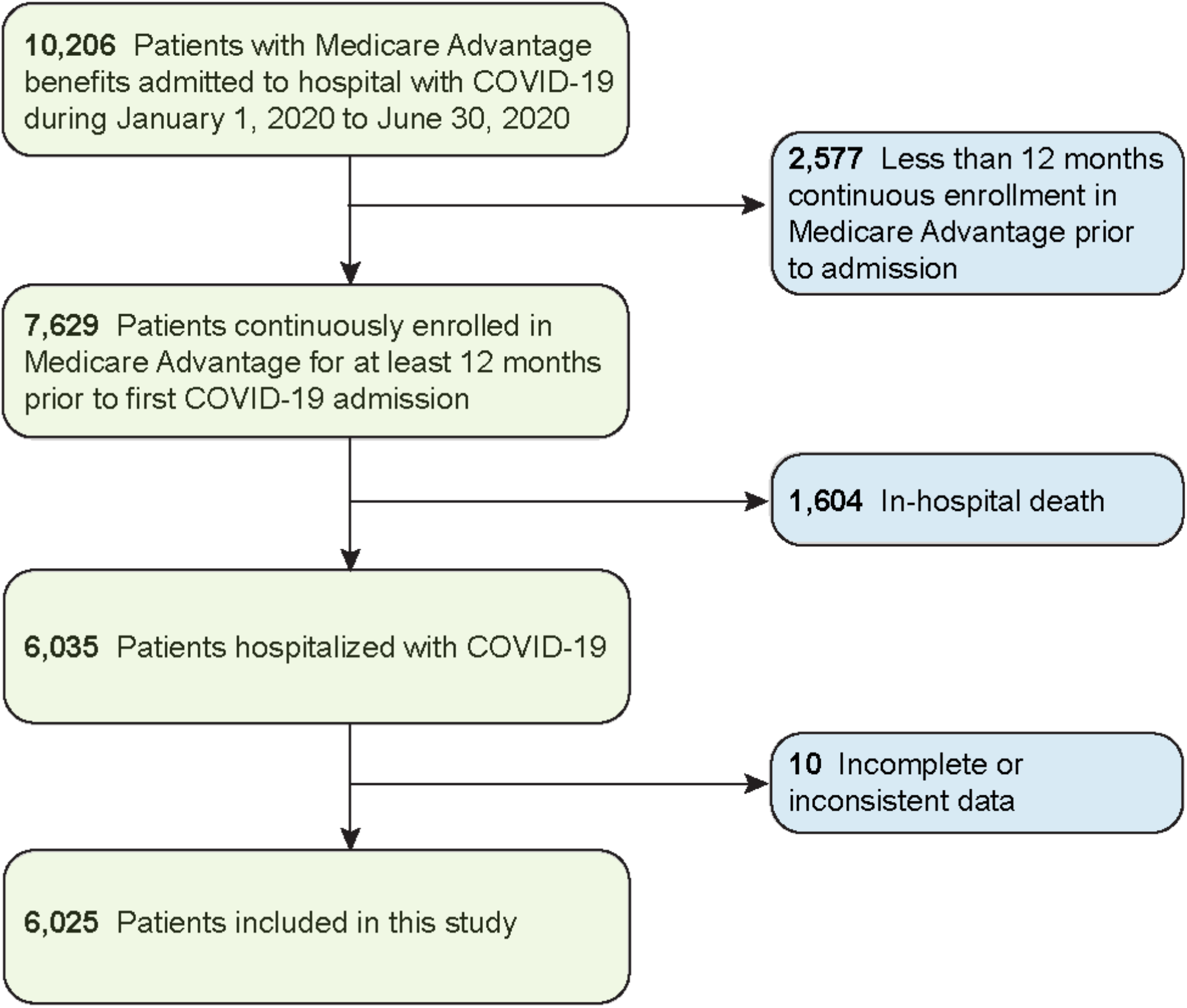
Selection of Study Patients.

**Table 1.** Characteristics of hospitalized COVID-19 Medicare Advantage members.

| Variable | Category | Value |
| --- | --- | --- |
| *Demographic / socioeconomic factors* |  |  |
| Gender | Female, No. (%) | 3,653 (61) |
| Age | Mean (SD) | 77 (11) |
| Race | White (non-Hispanic), No. (%) | 3,753 (62) |
|  | Black (non-Hispanic), No. (%) | 1,791 (30) |
|  | Hispanic / Latino, No. (%) | 191 (3) |
|  | Other, No. (%) | 140 (2) |
|  | Asian / Pacific Islander, No. (%) | 99 (2) |
|  | Unknown, No. (%) | 51 (1) |
| Residential location | Urban, No. (%) | 2,853 (47) |
|  | Suburban, No. (%) | 2,200 (37) |
|  | Rural, No. (%) | 972 (16) |
| Median income | Mean (SD) | \$63,027 (\$17,435) |
| *Plan and clinical characteristics* |  |  |
| C-SNP | Enrolled, No. (%) | 268 (4) |
| D-SNP | Enrolled, No. (%) | 332 (6) |
| I-SNP | Enrolled, No. (%) | 2,061 (34) |
| HCC count | 0 or 1, No. (%) | 1,460 (24) |
|  | 2 or 3, No. (%) | 1,991 (33) |
|  | 4 or 5, No. (%) | 1,182 (20) |
|  | 6 or more, No. (%) | 1,392 (23) |
| Chronic conditions | COPD, No. (%) | 1,450 (24) |
|  | Diabetes, No. (%) | 3,104 (52) |
|  | Hyperlipidemia, No. (%) | 3,499 (58) |
|  | Hypertension, No. (%) | 1,902 (32) |
| COVID-19 hospitalization (in days) | Mean LOS (SD) | 12.6 (11) |
| PIP | Low, ≤25%, No. (%) | 3,514 (58) |
|  | Moderate, 26%–50%, No. (%) | 1,784 (30) |
|  | High, >50%, No. (%) | 727 (12) |
| DDI | Minimal or minor, No. (%) | 3,110 (52) |
|  | Moderate, No. (%) | 983 (16) |
|  | Major, No. (%) | 1,542 (25) |
|  | Contraindicated, No. (%) | 390 (7) |

C-SNP: chronic conditions special needs plan (SNP); D-SNP: dual-eligible SNP; I-SNP: institutional SNP; HCCs: hierarchical conditions categories; LOS: length of stay; PIP: pharmacogenetic interaction probability; DDI: drug-drug interaction

Overall, 42% (2,511 of 6,025) of patients had a moderate or high PIP (Table 1), indicating that among these patients, there was a >25% chance that pharmacogenomic testing would reveal a clinically significant gene-drug or gene-drug-drug interaction. Further, 48% (2,915 of 6,025) had a moderate, major, or contraindicated DDI. Nearly one quarter (24%) of patients were prescribed metoprolol, a beta blocker associated with a minimum PIP of 46% (eTable 1). Other frequently prescribed drugs associated with moderate or high PIP were proton-pump inhibitors pantoprazole (12% of patients) and omeprazole (9%), and serotonin-reuptake inhibitors escitalopram (6%) and citalopram (3%) (eTable 1).

Patients were hospitalized for a mean of 13 days (SD, 11 days) (Table 1). Patients with moderate or high PIP were hospitalized for at least one day longer on average compared to those with low PIP, while LOS for patients with moderate, major, or contraindication DDI was similar to those with minimal or minor DDI (Figure 2 and eTable 2). In unadjusted analyses, the LOS for patients with moderate and high PIP were 9% (CI: 4%–15%; *P*<0.001) and 16% longer (CI: 8%–24%; *P*<0.001), respectively, compared to those with low PIP (eTable 3). Moreover, the LOS for male patients was 8% longer (CI: 3%–13%; *P*=0.001) compared to female patients, and for each one-year age increment, the expected LOS was 0.5% longer (CI: 0.3%–0.7%; *P*<0.001).

**Figure 2.**
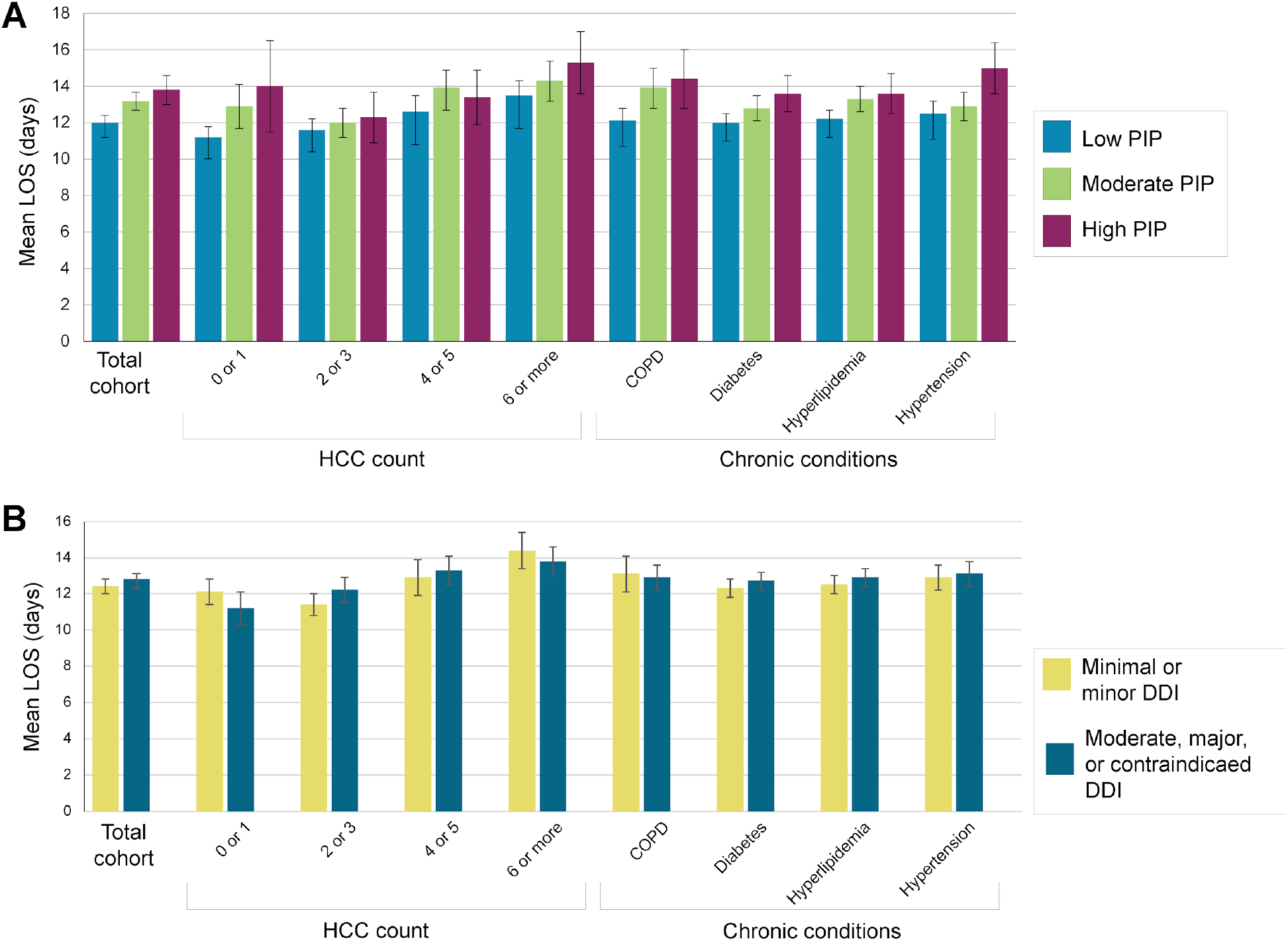
Length of stay by pharmacogenomic risk among Medicare Advantage members hospitalized with COVID-19. Average length of stay (LOS) in days among the entire cohort and subpopulations stratified by HCC count or chronic conditions was compared by (**A**) pharmacogenomic interaction probability (PIP) and (**B**) drug-drug interaction (DDI). Error bars show 95% confidence intervals. PIP: pharmacogenetic interaction probability; HCC: hierarchical condition categories; COPD: chronic obstructive pulmonary disease.

Among the entire cohort, adjusting for potential confounders did not attenuate the association between PIP and LOS (Table 2). Notably, across all patients, moderate and high PIP scores were significant predictors of LOS but RAF score was not (eTable 6). When patients were stratified by HCC count, PIP was significantly associated with LOS among all patient groups except those with 2 or 3 conditions (Table 2). In particular, among patients with 0 or 1, or 4 or 5 conditions, those with moderate PIP had 15% (CI: 4%–28%; *P*=0.007) and 13% (CI: 2%–25%; *P*=0.019) longer LOS, respectively, compared to those with low PIP. Moreover, among patients with 0 or 1, or 6 or more conditions, those with high PIP had 39% (CI: 15%–67%; *P*<0.001) and 16% (CI: 3%–31%; *P*=0.014) longer LOS, respectively, compared to those with low PIP.

**Table 2.** Expected length-of-stay ratios by pharmacogenetic risk and drug-drug interactions among patients by HCC count and chronic conditions.

| Population | Moderate PIP (26% to 50%)* |  | High PIP (>50%)* |  | Moderate, Major, or Contraindicated DDI** |  |
| --- | --- | --- | --- | --- | --- | --- |
|  | Rate ratio (95% CI) | P-value | Rate ratio (95% CI) | P-value | Rate ratio (95% CI) | P-value |
| Total cohort | 1.09 (1.04, 1.14) | < 0.001 | 1.16 (1.09, 1.24) | < 0.001 | 1.04 (1.00, 1.09) | 0.066 |
| HCC count |  |  |  |  |  |  |
| Any | 1.09 (1.04, 1.14) | < 0.001 | 1.16 (1.09, 1.24) | < 0.001 | 1.04 (1.00, 1.09) | 0.066 |
| 0 or 1 | 1.15 (1.04, 1.28) | 0.007 | 1.39 (1.15, 1.67) | < 0.001 | 0.91 (0.82, 1.00) | 0.045 |
| 2 or 3 | 1.03 (0.95, 1.11) | 0.522 | 1.08 (0.96, 1.21) | 0.204 | 1.10 (1.02, 1.18) | 0.010 |
| 4 or 5 | 1.13 (1.02, 1.25) | 0.019 | 1.13 (1.00, 1.29) | 0.057 | 1.07 (0.97, 1.17) | 0.179 |
| 6 or more | 1.08 (0.98, 1.18) | 0.112 | 1.16 (1.03, 1.31) | 0.014 | 1.01 (0.93, 1.11) | 0.786 |
| Chronic conditions |  |  |  |  |  |  |
| COPD | 1.13 (1.03, 1.24) | 0.009 | 1.18 (1.05, 1.34) | 0.006 | 1.01 (0.93, 1.10) | 0.796 |
| Diabetes | 1.05 (0.99, 1.12) | 0.119 | 1.15 (1.05, 1.25) | 0.002 | 1.07 (1.01, 1.13) | 0.031 |
| Hyperlipidemia | 1.08 (1.02, 1.15) | 0.013 | 1.12 (1.02, 1.22) | 0.014 | 1.05 (0.99, 1.11) | 0.096 |
| Hypertension | 1.03 (0.95, 1.12) | 0.478 | 1.22 (1.10, 1.35) | < 0.001 | 1.02 (0.95, 1.1) | 0.522 |

\*0-25% as baseline; \*\*minimal or minor as baseline
HCCs: hierarchical conditions categories; PIP: pharmacogenetic interaction probability; DDI: drug-drug interaction; COPD: chronic obstructive pulmonary disease. Significance was set at $P < 0.05$ .

Varying associations were observed for LOS, HCC count, and DDI. Among patients with 0 or 1 HCC, those with moderate or greater DDI had a 9% shorter LOS (CI: 0–18%; *P*=0.045) compared to those with minimal or minor DDI (Table 2). Among patients with 2 or 3 HCC, those with moderate or more severe DDI had 10% longer LOS (CI: 2–18%; *P*=0.010) compared to those with minimal or minor DDI. Lastly, across all HCC categories, enrollment in an I-SNP was associated with a 23%–34% shorter LOS compared to those not enrolled in I-SNP (eTables 6– 10).

To explore the impact of individual comorbidities on LOS, patients were stratified by four pre-existing chronic conditions: COPD, diabetes, hyperlipidemia, and hypertension. Patients with high PIP most frequently had hyperlipidemia (eTable 11). Within each chronic condition subpopulation (controlled for the other chronic conditions), patients with high PIP were hospitalized 12%–22% longer compared to those with low PIP (Table 2). In particular, among patients with hypertension, those with high PIP were hospitalized 2.5 days longer than those with low PIP (Table 3). Moreover, patients diagnosed with hyperlipidemia or COPD who had moderate PIP had longer LOS by 8% (CI: 2%–15%; *P*=0.013) and 13% (CI: 3%–24%; *P*=0.009), respectively, compared to those with low PIP (Table 2).

**Table 3.** Distribution of length of stay among patients hospitalized with COVID-19 by chronic condition and pharmacogenomic or drug-drug interaction risk.

| **Chronic condition** | **Total subpopulation, mean LOS (95% CI)** | **Mean LOS (95% CI) by PIP** |  |  | **Mean LOS (95% CI) by DDI** |  |
| --- | --- | --- | --- | --- | --- | --- |
|  |  | **Low (≤25%)** | **Moderate (26–50%)** | **High (>50%)** | **Minimal or minor** | **Moderate, major, or contraindicated** |
| COPD | 13.0 (12.4, 13.6) | 12.1 (11.4, 12.8) | 13.9 (12.8, 15) | 14.4 (12.8, 16) | 13.1 (12.1, 14.1) | 12.9 (12.2, 13.6) |
| Diabetes | 12.5 (12.1, 12.9) | 12 (11.5, 12.5) | 12.8 (12.1, 13.5) | 13.6 (12.6, 14.6) | 12.3 (11.8, 12.8) | 12.7 (12.2, 13.2) |
| Hyperlipidemia | 12.7 (12.3, 13.1) | 12.2 (11.7, 12.7) | 13.3 (12.6, 14) | 13.6 (12.5, 14.7) | 12.5 (12, 13) | 12.9 (12.4, 13.4) |
| Hypertension | 13.0 (12.5, 13.5) | 12.5 (11.8, 13.2) | 12.9 (12.1, 13.7) | 15 (13.6, 16.4) | 12.9 (12.2, 13.6) | 13.1 (12.4, 13.8) |

LOS: length of stay; PIP: pharmacogenomic interaction probability; DDI: drug-drug interaction.

When considering the potential impact of DDI on LOS, we found that patients diagnosed with diabetes and at least moderate or more severe DDI had 7% longer LOS (CI: 1%–13%; *P*=0.031) compared to those with minimal or minor DDI (Table 2). Lastly, patients in each chronic condition group who were enrolled in I-SNP had a 25%–29% shorter LOS than those not enrolled (eTables 12–15).

## Discussion

In this study, we showed that greater pharmacogenomic risk was associated with longer LOS among Medicare Advantage members hospitalized with COVID-19. In particular, high PIP patients, who have a 50% or greater likelihood of harboring a genetics-driven atypical response to current medications, were hospitalized for almost two days longer than those with low PIP. Given the known associations between ADRs and LOS,^8^ our findings suggest that incorporating pharmacogenomic testing into clinical management of high-risk COVID-19 patients may shorten hospital LOS. Critically, to potentially reduce LOS, testing would have to occur prior to admission to allow for testing results to be incorporated into a patient’s medication management.

In a novel pandemic such as the COVID-19 crisis, the sudden and intense demands on hospitals present a clear need to reduce patient LOS. Should proactive pharmacogenomics-informed medication management for COVID-19 patients successfully reduce LOS, even non-COVID patients could benefit by reducing the burden on overwhelmed healthcare systems that have delayed some patient care.^16,33–35^ In addition, pharmacogenomic intervention could reduce COVID-19 healthcare costs. Estimating the potential cost savings from a reduction in LOS is challenging; however, recent data show that the average Medicare payment per COVID-19 hospitalization is $23,587,^18^ which is $8,688 higher than the average Medicare Advantage hospitalization ($14,900).^36^ Considering the higher cost associated with complex COVID-19 patients, pharmacogenomic-informed medication management of Medicare beneficiaries could result in substantial savings for patients, hospitals, and the Medicare program.

Overall, we found that almost half of Medicare Advantage members hospitalized with COVID-19 could benefit from improved medication management, as 42% had a moderate or high PIP and 48% had a moderate, major, or contraindicated DDI for current medications.^9–11^ Further, despite mandates to flag DDIs in electronic health records and pharmacy systems, we found that one-third of our cohort was prescribed medications for which risks likely exceeded benefits. Thus, comprehensive medication management is underutilized among Medicare members, leaving many at risk for ADRs. Incorporating pharmacy expertise into the clinical management of patients could reduce this risk as pharmacist participation has been shown to reduce ADRs, LOS, and mortality.^39^

Several findings in our subpopulation analyses warrant further study. When patients were stratified by number of chronic conditions, patients with 0 or 1 HCC and high PIP had a greater LOS increase than those with 6 or more HCC (39% vs 16% increase), which may be due to uncontrolled confounders such as severity of COVID-19 infection. In addition, we did not observe a significant association between PIP and LOS among patients with 2 or 3 chronic conditions despite finding a significant association for all other HCC counts. A larger sample size may reveal significant findings for those with 2 or 3 HCC and possibly identify additional confounders. In addition, the finding that patients with 0 or 1 HCC and more severe DDI had shorter LOS compared to those with less severe DDI requires further study that examines the specific medications used by those patients. Separately, for each specific chronic condition (COPD, diabetes, hyperlipidemia, and hypertension), we found patients with high PIP had significantly longer LOS compared to those with low PIP. Further research may explore the impact of pharmacogenomic risk and ADRs among COVID-19 patients with these comorbidities.

Finally, in both subpopulation analyses, I-SNP enrollment was associated with shorter LOS. This may be a result of potential ADRs receiving more attention in institutional settings, or conversely, an effect of more expedient discharge due to enhanced care coordination with institutional settings.

LOS is a long-standing concern for healthcare organization performance, patient outcomes, and expenditures.^40,41,42^ Although this study focused on COVID-19, population-based pharmacogenomic testing that improves medication management could broadly reduce ADRs, which are reported to cost the US $528 billion per year, exceeding the costs associated with any major disease or the actual drugs prescribed.^43^ A one-time test could aid medication optimization and potentially reduce healthcare costs throughout each patient’s life. In addition, future research should further explore the potential for PIP to predict LOS, as moderate and high PIP were both significant predictors of LOS whereas RAF score, a metric used by CMS for benchmarking expenditures and allocating payments to healthcare providers, was not (eTable 6).

## Limitations

Limitations to this study include that although this study showed an association between PIP and increased LOS, it was not a study demonstrating that increased LOS among COVID-19 patients was a direct result of adverse drug events that could have been prevented by proactive pharmacogenomic testing. Future studies with large, controlled cohorts are needed to demonstrate the impact of pharmacogenomics-guided medication management on hospital LOS. In addition, the analyses did not account for some potential confounders such as severity of COVID-19, medication regimen complexity (including the types and counts of prescribed drugs, medication dosage or frequency), or patient kidney or hepatic function. A general limitation of observational studies using claims data is that the presence of a code does not guarantee that a patient has been diagnosed or has received or taken a prescribed medication. Finally, our findings may have been affected by a survivor bias arising from the exclusion of patients who died during the study period.

## Conclusions

Among otherwise risk-matched patients with COVID-19, moderate or high pharmacogenomic risk was associated with longer LOS. Addressing pharmacogenomic risk prior to COVID-19 hospitalization or other serious disease may reduce LOS, decrease healthcare costs, and improve risk predictions in Medicare members, especially those with COPD, diabetes, hyperlipidemia, or hypertension.

## Supporting information

Supplemental eTables

## Data Availability

The data analyzed in this study was obtained from UnitedHealth Group Clinical Discovery Portal. The data are proprietary and are not available for public use but, under certain conditions, may be made available to editors and their approved auditors under a data-use agreement to confirm the findings of the current study. Further inquiries can be directed to Lauren Mihajlov.

## List of abbreviations

COVID-19: Coronavirus-induced disease 2019
CPIC: Clinical Pharmacogenetics Implementation Consortium
DDI: Drug-drug interaction
FDA: U.S. Food and Drug Administration
HCC: hierarchical conditions categories
LOS: Length of stay
PIP: Pharmacogenetic interaction probability
RAF: Risk adjustment factor
SNP: Special needs plan

## Acknowledgements

Taryn Hall had full access to all the data in the study and takes responsibility for the integrity of the data and the accuracy of the data analysis.

The authors thank Kathy Tzeng of UnitedHealth Group (UHG) for the initial development of this project, Nicolas Moyer of Invitae for assisting with analyses, and Elaine Preimesberger of UHG and Melinda Tran of Invitae for their review and feedback on this draft.

Authors KA, CM, CS, and SR are employees of genetic testing company Invitae. Authors KVT, GOM, JR, BM, YG, ETE, and TOH are employees of UHG.

