## Supplemental eTables for "Pharmacogenomic and drug interaction risk associations with hospital length of stay among Medicare Advantage members with COVID-19"

**Supplemental Tables**

**eTable 1:** Frequently prescribed drugs with moderate or high PIP among Medicare Advantage members hospitalized with COVID-19
**eTable 2:** Distribution of length of stay among Medicare Advantage members hospitalized with COVID-19 by pharmacogenomic risk and drug-drug interaction
**eTable 3:** Length-of-stay ratios of covariates in the baseline model
**eTable 4:** Distribution of patients by HCC count subpopulation, PIP, and DDI category
**eTable 5:** Distribution of length of stay among Medicare Advantage members hospitalized with COVID-19 by HCC count and pharmacogenomic or drug-drug interaction risk

**eTables 6–10: Summary Tables for HCC Count Subpopulation Models**
**eTable 6:** Length-of-stay ratios in the model for all Medicare Advantage members hospitalized with COVID-19
**eTable 7:** Expected LOS ratios in the model for patients with zero or one HCC
**eTable 8:** Expected LOS ratios in the model for patients with two or three HCC
**eTable 9:** Expected LOS ratios in the model for patients with four or five HCC
**eTable 10:** Expected LOS ratios in the model for patients with six or more HCC

**eTable 11:** Distribution of patients by chronic condition subpopulation, pharmacogenomic risk, and drug-drug interaction risk

**eTables 12–15: Summary Tables for Chronic Conditions Subpopulation Models**
**eTable 12:** Expected LOS ratios in the model for patients with COPD
**eTable 13:** Expected LOS ratios in the model for patients with diabetes
**eTable 14:** Expected LOS ratios in the model for patients with hyperlipidemia
**eTable 15:** Expected LOS ratios in the model for patients with hypertension

**eTable 1. Frequently prescribed drugs with moderate or high PIP among Medicare Advantage members hospitalized with COVID-19**

| Generic Name | Drug Class | Minimum patient PIP associated with drug | Frequency, No. (%) |
|---|---|---|---|
| Metoprolol | Beta blockers | 46% | 1,422 (24%) |
| Pantoprazole | Proton-pump inhibitors | 29% | 730 (12%) |
| Omeprazole | Proton-pump inhibitors | 30% | 571 (9%) |
| Escitalopram | Selective serotonin-reuptake Inhibitors | 33% | 365 (6%) |
| Citalopram | Selective serotonin-reuptake Inhibitors | 32% | 207 (3%) |

PIP, pharmacogenetic interaction probability

### eTable 2. Distribution of length of stay among Medicare Advantage members hospitalized with COVID-19 by pharmacogenomic risk and drug-drug interaction

| Medication risk measure | Pharmacogenomic risk category | LOS (days), Mean (95% CI) | LOS (days) Median |
|---|---|---|---|
| PIP | Low (≤25%) | 12.0 (11.6–12.4) | 8 |
| | Moderate (26%–50%) | 13.2 (12.7–13.7) | 9 |
| | High (>50%) | 13.8 (13-14.6) | 9 |
| DDI | Minimal or minor | 12.4 (12–12.8) | 8 |
| | Moderate, major, or contraindicated | 12.7 (12.3–13.1) | 8 |

The average LOS (in days) for each unique DDI severity category were minimal: 12.0 for minimal DDI, 12.5 for minor DDI, 13.1 for moderate DDI, 12.6 for major DDI, and 12.6 for contraindicated DDI.
LOS: length of stay; PIP: pharmacogenetic interaction probability; DDI: drug-drug interaction

### eTable 3. Length-of-stay ratios of covariates in the baseline model

| Variable | Rate Ratio (95% CI) | *P*-value |
|---|---|---|
| Moderate PIP (26% – 50%)[a] | 1.09 (1.04, 1.15) | <0.001 |
| High PIP (> 50%)[a] | 1.16 (1.08, 1.24) | <0.001 |
| Moderate, major, or contraindicated DDI[b] | 1.02 (0.98, 1.06) | 0.384 |
| Age | 1.005 (1.003, 1.007) | <0.001 |
| Gender[c] | 1.08 (1.03, 1.13) | 0.001 |

[a]Low PPI (0-25%) as baseline; [b]Minimal or minor DDI as baseline; [c]female as baseline.
PIP: pharmacogenetic interaction probability; DDI: drug-drug interaction

### eTable 4. Distribution of patients by HCC count subpopulation, pharmacogenomic risk, and drug-drug interaction categories

| HCC Count | Patients, No. (%) | No. of patients by PIP | | | No. of patients by DDI | |
|---|---|---|---|---|---|---|
| | | Low (≤25%) | Moderate (26%–50%) | High (>50%) | Minimal or minor | Moderate, major, or contraindicated |
| 0 or 1 | 1,460 (24.2) | 989 | 377 | 94 | 954 | 506 |
| 2 or 3 | 1,991 (33.1) | 1,245 | 542 | 204 | 1,083 | 908 |
| 4 or 5 | 1,182 (19.6) | 597 | 388 | 197 | 519 | 663 |
| 6 or more | 1,392 (23.1) | 683 | 477 | 232 | 554 | 838 |

HCC: hierarchical conditions categories; PIP: pharmacogenomic interaction probability; DDI: drug-drug interaction.

**eTable 5. Distribution of length of stay among Medicare Advantage members hospitalized with COVID-19 by HCC count and pharmacogenomic or drug-drug interaction risk**

| HCC Count | Mean LOS | LOS by PIP, mean (95% CI) | | | LOS by DDI, mean (95% CI) | |
|---|---|---|---|---|---|---|
| | | ≤ 25% | 26–50% | >50% | Minimal or minor | Moderate, major, or contraindicated |
| Any | 12.6 | 12.0 (11.6–12.4) | 13.2 (12.7–13.7) | 13.8 (13–14.6) | 12.4 (12–12.8) | 12.8 (12.3–13.1) |
| 0 or 1 | 11.8 | 11.2 (10.6–11.8) | 12.9 (11.7–14.1) | 14.0 (11.5–16.5) | 12.1 (11.4–12.8) | 11.2 (10.3–12.1) |
| 2 or 3 | 11.8 | 11.6 (11–12.2) | 12.0 (11.2–12.8) | 12.3 (10.9–13.7) | 11.4 (10.8–12) | 12.2 (11.5–12.9) |
| 4 or 5 | 13.1 | 12.6 (11.7–13.5) | 13.9 (12.7–14.9) | 13.4 (11.9–14.9) | 12.9 (11.9–13.9) | 13.3 (12.5–14.1) |
| 6 or more | 14.1 | 13.5 (12.5–14.3) | 14.3 (13.2–15.4) | 15.3 (13.6–17) | 14.4 (13.4–15.4) | 13.8 (13–14.6) |

HCC: hierarchical conditions categories; PIP: pharmacogenomic interaction probability; DDI: drug-drug interaction

**eTable 6. Length-of-stay ratios in the model for all Medicare Advantage members hospitalized with COVID-19**

| Variable | Description | Ratio | 95% C.I. | *P*-value |
|---|---|---|---|---|
| (Intercept) | Intercept | 7.12 | (5.41, 9.37) | < 0.001 |
| Moderate PIP[a] | 26 to 50% | 1.09 | (1.04, 1.14) | < 0.001 |
| High PIP[a] | > 50% | 1.16 | (1.09, 1.24) | < 0.001 |
| DDI Category 1[b] | Moderate, major, or contraindicated | 1.04 | (1.00, 1.09) | 0.066 |
| Age | Age of a patient (2019) | 1.007 | (1.004, 1.009) | < 0.001 |
| Income | Median household income per ZCTA level (standardized) | 1.06 | (1.04, 1.09) | < 0.001 |
| RAF Score | Risk adjustment factor score | 1.02 | (1.00, 1.04) | 0.108 |
| D-SNP | Enrolled for at least one month in the dual - eligible special needs plan | 0.93 | (0.85, 1.02) | 0.136 |
| I-SNP | Enrolled for at least one month in the institutional special needs plan | 0.73 | (0.70, 0.77) | < 0.001 |
| Gender[c] | Gender of a patient | 1.05 | (1.00, 1.10) | 0.027 |
| Race Code 1[d] | White (non-Hispanic) | 0.97 | (0.77, 1.21) | 0.769 |
| Race Code 2[d] | Black (non-Hispanic) | 0.97 | (0.77, 1.22) | 0.806 |
| Race Code 3[d] | Other | 1.03 | (0.79, 1.33) | 0.845 |
| Race Code 4[d] | Asian/Pacific Islander | 0.89 | (0.68, 1.18) | 0.426 |
| Race Code 5[d] | Hispanic/Latino | 0.90 | (0.70, 1.16) | 0.430 |
| HCC009 | Lung and other severe cancers | 0.76 | (0.64, 0.90) | 0.002 |
| HCC017 | Diabetes with acute complications | 0.88 | (0.74, 1.05) | 0.160 |

| | | | | |
|---|---|---|---|---|
| HCC023 | Other significant endocrine and metabolic disorders | 0.92 | (0.84, 1.01) | 0.079 |
| HCC027 | End-stage liver disease | 1.38 | (1.04, 1.82) | 0.026 |
| HCC033 | Intestinal obstruction/perforation | 1.05 | (0.93, 1.19) | 0.443 |
| HCC039 | Bone/joint/muscle infections/necrosis | 1.14 | (1.00, 1.30) | 0.051 |
| HCC046 | Severe hematological disorders | 0.73 | (0.57, 0.94) | 0.013 |
| HCC047 | Disorders of immunity | 0.92 | (0.79, 1.07) | 0.269 |
| HCC048 | Coagulation defects and other specified hematological disorders | 1.06 | (0.97, 1.15) | 0.192 |
| HCC054 | Substance use with psychotic complications | 1.27 | (0.92, 1.74) | 0.147 |
| HCC055 | Substance use disorder, moderate/severe or substance use with complications | 1.08 | (0.95, 1.23) | 0.234 |
| HCC057 | Schizophrenia | 1.13 | (1.02, 1.25) | 0.018 |
| HCC072 | Spinal cord disorders/injuries | 1.18 | (0.97, 1.44) | 0.106 |
| HCC073 | Amyotrophic lateral sclerosis and other motor neuron disease | 1.28 | (0.71, 2.30) | 0.416 |
| HCC077 | Multiple sclerosis | 1.41 | (1.20, 1.66) | < 0.001 |
| HCC082 | Respirator dependence/ tracheostomy status | 1.22 | (0.96, 1.56) | 0.101 |
| HCC084 | Cardio-respiratory failure and shock | 1.07 | (0.98, 1.16) | 0.124 |
| HCC099 | Cerebral hemorrhage | 1.09 | (0.91, 1.31) | 0.324 |
| HCC103 | Hemiplegia/hemiparesis | 1.06 | (0.97, 1.15) | 0.195 |
| HCC104 | Monoplegia, other paralytic syndromes | 0.63 | (0.43, 0.94) | 0.022 |
| HCC107 | Vascular disease with complications | 1.10 | (0.99, 1.23) | 0.088 |

| | | | | |
|---|---|---|---|---|
| HCC112 | Fibrosis of lung and other chronic lung disorders | 0.69 | (0.54, 0.89) | 0.004 |
| HCC114 | Aspiration and specified bacterial pneumonias | 1.02 | (0.90, 1.15) | 0.758 |
| HCC115 | Pneumococcal pneumonia, empyema, lung abscess | 0.91 | (0.80, 1.05) | 0.191 |
| HCC134 | Dialysis status | 0.91 | (0.80, 1.05) | 0.194 |
| HCC135 | Acute renal failure | 1.01 | (0.95, 1.09) | 0.684 |
| HCC137 | Chronic kidney disease, severe (stage 4) | 0.83 | (0.69, 1.00) | 0.046 |
| HCC161 | Chronic ulcer of skin, except pressure | 1.07 | (0.98, 1.17) | 0.144 |
| HCC169 | Vertebral fractures without spinal cord injury | 1.13 | (0.96, 1.32) | 0.149 |
| HCC170 | Hip fracture/dislocation | 1.23 | (1.09, 1.38) | 0.001 |
| HCC173 | Traumatic amputations and complications | 1.16 | (0.94, 1.43) | 0.171 |

[a]0-25% PPI as baseline; [b]minimal or minor DDI as baseline; [c]female as baseline; [d]unknown race as baseline
Significance was set at $P<0.05$. HCC: hierarchical conditions categories. PIP: pharmacogenomic interaction probability; DDI: drug-drug interaction; D-SNP: dual special needs plan; I-SNP: institutional special needs plan.

**eTable 7. Expected length-of-stay ratios in the model for patients with zero or one HCC**

| Variable | Description | Ratio | 95% C.I. | *P*-value |
|---|---|---|---|---|
| (Intercept) | Intercept | 4.93 | (2.91, 8.34) | <0.001 |
| Moderate PIP[a] | 26 to 50% | 1.15 | (1.04, 1.28) | 0.007 |
| High PIP[a] | >50% | 1.39 | (1.15, 1.67) | <0.001 |
| DDI Category 1[b] | Moderate, major, or contraindicated | 0.91 | (0.82, 1.00) | 0.045 |
| Age | Age of a patient (2019) | 1.015 | (1.010, 1.020) | <0.001 |
| Income | Median household income per ZCTA level (standardized) | 1.03 | (0.99, 1.08) | 0.188 |
| C-SNP | Enrolled for at least one month in the chronic or disabling condition special needs plan | 0.89 | (0.68, 1.15) | 0.372 |
| D-SNP | Enrolled for at least one month in the dual - eligible special needs plan | 0.74 | (0.58, 0.94) | 0.012 |
| I-SNP | Enrolled for at least one month in the institutional special needs plan | 0.75 | (0.67, 0.84) | <0.001 |
| Gender[c] | Gender of a patient | 1.08 | (0.98, 1.19) | 0.107 |
| Race Code 1[d] | White (non-Hispanic) | 0.73 | (0.50, 1.06) | 0.101 |
| Race Code 2[d] | Black (non-Hispanic) | 0.69 | (0.47, 1.01) | 0.059 |
| Race Code 3[d] | Other | 0.57 | (0.36, 0.88) | 0.012 |
| Race Code 4[d] | Asian/Pacific Islander | 0.73 | (0.47, 1.14) | 0.166 |
| Race Code 5[d] | Hispanic/Latino | 0.61 | (0.40, 0.93) | 0.022 |
| HCC010 | Lymphoma and other cancers | 1.36 | (0.72, 2.56) | 0.338 |
| HCC012 | Breast, prostate, and other cancers and tumors | 0.86 | (0.63, 1.17) | 0.333 |

| | | | | |
|---|---|---|---|---|
| HCC022 | Morbid obesity | 0.87 | (0.59, 1.30) | 0.506 |
| HCC040 | Rheumatoid arthritis and inflammatory connective tissue disease | 1.52 | (1.10, 2.10) | 0.012 |
| HCC057 | Schizophrenia | 1.50 | (0.97, 2.33) | 0.071 |
| HCC058 | Reactive and unspecified psychosis | 1.40 | (1.08, 1.82) | 0.011 |
| HCC077 | Multiple sclerosis | 1.45 | (0.79, 2.63) | 0.227 |
| HCC079 | Seizure disorders and convulsions | 0.64 | (0.41, 1.00) | 0.052 |
| HCC085 | Congestive heart failure | 0.88 | (0.65, 1.20) | 0.422 |
| HCC115 | Pneumococcal pneumonia, empyema, lung abscess | 2.62 | (1.33, 5.17) | 0.005 |
| HCC124 | Exudative macular degeneration | 1.15 | (0.67, 1.95) | 0.616 |

[a]0-25% PIP as baseline; [b]minimal or minor DDI as baseline; [c]female as baseline; [d]unknown race as baseline
Significance was set at $P<0.05$. HCC: hierarchical conditions categories. PIP: pharmacogenomic interaction probability; DDI: drug-drug interaction; C-SNP: chronic condition special needs plan; D-SNP: dual special needs plan; I-SNP: institutional special needs plan.

**eTable 8. Expected length-of-stay ratios in the model for patients with two or three HCC**

| Variable | Description | Ratio | 95% C.I. | *P*-value |
|---|---|---|---|---|
| (Intercept) | Intercept | 6.25 | (3.91, 10.01) | <0.001 |
| Moderate PIP[a] | 26% to 50% | 1.03 | (0.95, 1.11) | 0.522 |
| High PIP[a] | >50% | 1.08 | (0.96, 1.21) | 0.204 |
| DDI Category 1[b] | Moderate, major, or contraindicated | 1.10 | (1.02, 1.18) | 0.010 |
| Age | Age of a patient (2019) | 1.004 | (1.000, 1.007) | 0.028 |
| Income | Median household income per ZCTA level (standardized) | 1.07 | (1.03, 1.11) | <0.001 |
| I-SNP | Enrolled for at least one month in the institutional special needs plan | 0.77 | (0.72, 0.83) | <0.001 |
| Race Code 1[c] | White (non-Hispanic) | 1.42 | (0.94, 2.14) | 0.098 |
| Race Code 2[c] | Black (non-Hispanic) | 1.46 | (0.97, 2.21) | 0.073 |
| Race Code 3[c] | Other | 1.74 | (1.10, 2.76) | 0.018 |
| Race Code 4[c] | Asian/Pacific Islander | 1.10 | (0.66, 1.83) | 0.720 |
| Race Code 5[c] | Hispanic/Latino | 1.54 | (0.98, 2.44) | 0.061 |
| HCC018 | Diabetes with chronic complications | 0.92 | (0.86, 1.00) | 0.043 |
| HCC023 | Other significant endocrine and metabolic disorders | 0.81 | (0.66, 1.00) | 0.045 |
| HCC054 | Substance use with psychotic complications | 1.66 | (0.91, 3.03) | 0.099 |
| HCC058 | Reactive and unspecified psychosis | 1.09 | (0.99, 1.21) | 0.084 |
| HCC072 | Spinal cord disorders/injuries | 1.67 | (1.10, 2.52) | 0.015 |
| HCC084 | Cardio-respiratory failure and shock | 1.40 | (1.03, 1.90) | 0.033 |

| | | | | |
|---|---|---|---|---|
| HCC106 | Atherosclerosis of extremity with ulceration or gangrene | 1.51 | (1.05, 2.17) | 0.025 |
| HCC112 | Fibrosis of lung and other chronic lung disorders | 0.46 | (0.27, 0.78) | 0.004 |
| HCC169 | Vertebral fractures without spinal cord injury | 1.25 | (0.90, 1.72) | 0.177 |
| HCC170 | Hip fracture/dislocation | 1.45 | (1.13, 1.85) | 0.003 |

[a]0-25% PIP as baseline; [b]minimal or minor DDI as baseline; [c]unknown race as baseline. Significance was set at $P<0.05$.
HCC: hierarchical condition categories; PIP: pharmacogenomic interaction probability; DDI: drug-drug interaction; I-SNP: institutional special needs plan.

**eTable 9. Expected length-of-stay ratios in the model for patients with four or five HCC**

| Variable | Description | Ratio | 95% C.I. | *P*-value |
|---|---|---|---|---|
| (Intercept) | Intercept | 12.97 | (11.92, 14.11) | <0.001 |
| Moderate PIP[a] | 26 to 50% | 1.13 | (1.02, 1.25) | 0.019 |
| High PIP[a] | >50% | 1.13 | (1.00, 1.29) | 0.057 |
| DDI Category 1[b] | Moderate, major, or contraindicated | 1.07 | (0.97, 1.17) | 0.179 |
| Income | Median household income per ZCTA level (standardized) | 1.05 | (1.01, 1.10) | 0.021 |
| I-SNP | Enrolled for at least one month in the institutional special needs plan | 0.72 | (0.65, 0.79) | <0.001 |
| HCC09 | Lung and other severe cancers | 0.59 | (0.42, 0.83) | 0.002 |
| HCC039 | Bone/joint/muscle infections/necrosis | 1.65 | (1.22, 2.22) | 0.001 |
| HCC072 | Spinal cord disorders/injuries | 1.44 | (0.92, 2.28) | 0.114 |
| HCC077 | Multiple sclerosis | 1.45 | (1.09, 1.92) | 0.010 |
| HCC173 | Traumatic amputations and complications | 1.35 | (0.85, 2.13) | 0.199 |

[a]0-25% PIP as baseline; [b]minimal or minor DDI as baseline. Significance was set at $P<0.05$.
HCC: hierarchicial conditions categories; PIP: pharmacogenomic interaction probability; DDI: drug-drug interaction; I-SNP: institutional special needs plan.

**eTable 10. Expected length-of-stay ratios in the model for patients with six or more chronic conditions**

| Variable | Description | Ratio | 95% C.I. | *P*-value |
|---|---|---|---|---|
| (Intercept) | Intercept | 12.37 | (10.54, 14.51) | <0.001 |
| Moderate PIP[a] | 26% to 50% | 1.08 | (0.98, 1.18) | 0.112 |
| High PIP[a] | >50% | 1.16 | (1.03, 1.31) | 0.014 |
| DDI Category 1[b] | Moderate, major, or contraindicated | 1.01 | (0.93, 1.11) | 0.786 |
| Income | Median household income per ZCTA level (standardized) | 1.08 | (1.03, 1.12) | <0.001 |
| RAF Score | Risk adjustment factor score | 1.02 | (0.99, 1.05) | 0.239 |
| I-SNP | Enrolled for at least one month in the institutional special needs plan | 0.66 | (0.59, 0.73) | <0.001 |
| Gender[c] | Gender of a patient | 1.11 | (1.01, 1.20) | 0.022 |
| HCC008 | Metastatic cancer and acute leukemia | 1.26 | (0.99, 1.58) | 0.056 |
| HCC009 | Lung and other severe cancers | 0.83 | (0.65, 1.06) | 0.134 |
| HCC012 | Breast, prostate, and other cancers and tumors | 1.20 | (1.02, 1.41) | 0.025 |
| HCC023 | Other significant endocrine and metabolic disorders | 0.91 | (0.81, 1.03) | 0.135 |
| HCC027 | End-stage liver disease | 1.34 | (0.98, 1.84) | 0.069 |
| HCC028 | Cirrhosis of liver | 0.75 | (0.56, 1.02) | 0.064 |
| HCC046 | Severe hematological disorders | 0.54 | (0.38, 0.76) | <0.001 |
| HCC054 | Substance use with psychotic complications | 1.35 | (0.88, 2.07) | 0.173 |
| HCC055 | Substance use disorder, moderate/severe or substance use with complications | 1.11 | (0.95, 1.30) | 0.194 |

| | | | | |
|---|---|---|---|---|
| HCC077 | Multiple sclerosis | 1.37 | (1.06, 1.78) | 0.018 |
| HCC099 | Cerebral hemorrhage | 1.30 | (1.05, 1.60) | 0.015 |
| HCC108 | Vascular disease | 0.89 | (0.82, 0.97) | 0.009 |
| HCC112 | Fibrosis of lung and other chronic lung disorders | 0.66 | (0.46, 0.95) | 0.025 |
| HCC114 | Aspiration and specified bacterial pneumonias | 1.05 | (0.92, 1.19) | 0.479 |
| HCC124 | Exudative macular degeneration | 1.23 | (0.98, 1.55) | 0.073 |
| HCC134 | Dialysis status | 0.86 | (0.74, 1.01) | 0.075 |
| HCC135 | Acute renal failure | 1.06 | (0.96, 1.16) | 0.252 |
| HCC158 | Pressure ulcer of skin with full thickness skin loss | 1.15 | (0.99, 1.34) | 0.069 |
| HCC170 | Hip fracture/dislocation | 1.12 | (0.95, 1.33) | 0.167 |

[a]0-25% PIP as baseline; [b]minimal or minor DDI as baseline; [c]female as baseline
Significance was set at *P*<0.05. PIP: pharmacogenomic interaction probability; DDI: drug-drug interaction; RAF: risk adjustment factor; I-SNP: institutional special needs plan.

**eTable 11. Distribution of patients by chronic condition subpopulation, pharmacogenomic risk, and drug-drug interaction risk**

| Condition Group | Patient Count (%) | No. of patients by PIP | | | No. of patients by DDI | |
|---|---|---|---|---|---|---|
| | | ≤ 25% | 26% to 50% | > 50% | Minimal or minor | Moderate, major, or contraindicated |
| Hypertension | 1,902 (31.6) | 984 | 620 | 298 | 887 | 1,015 |
| Hyperlipidemia | 3,499 (58.1) | 1964 | 1078 | 457 | 1779 | 1,720 |
| Diabetes | 3,104 (51.5) | 1675 | 983 | 446 | 1525 | 1,579 |
| COPD | 1,450 (24.1) | 804 | 440 | 206 | 598 | 852 |

**eTable 12. Expected length-of-stay ratios in the model for patients with chronic obstructive pulmonary disease**

| Variable | Description | Ratio | 95% C.I. | *P*-value |
|---|---|---|---|---|
| (Intercept) | Intercept | 5.31 | (2.76, 10.24) | <0.001 |
| Moderate PIP[a] | 26% to 50% | 1.13 | (1.03, 1.24) | 0.009 |
| High PIP[a] | >50% | 1.18 | (1.05, 1.34) | 0.006 |
| DDI Category 1[b] | Moderate, major, or contraindicated | 1.01 | (0.93, 1.10) | 0.796 |
| Hypertension | Hypertension flag | 1.05 | (0.96, 1.15) | 0.318 |
| Hyperlipidemia | Hyperlipidemia flag | 1.07 | (0.98, 1.16) | 0.122 |
| Diabetes | Diabetes flag | 0.95 | (0.87, 1.04) | 0.248 |
| Age | Age of a patient (2019) | 1.006 | (1.001, 1.010) | 0.010 |
| Income | Median household income per ZCTA level (standardized) | 1.04 | (1.00, 1.09) | 0.058 |
| RAF Score | Risk adjustment factor score | 1.01 | (0.98, 1.04) | 0.603 |
| Gender[c] | Gender of a patient | 1.10 | (1.01, 1.20) | 0.028 |
| Race Code 1[d] | White (non-Hispanic) | 1.40 | (0.80, 2.45) | 0.233 |
| Race Code 2[d] | Black (non-Hispanic) | 1.32 | (0.76, 2.32) | 0.325 |
| Race Code 3[d] | Other | 1.37 | (0.71, 2.61) | 0.346 |
| Race Code 4[d] | Asian/Pacific Islander | 1.14 | (0.58, 2.21) | 0.705 |
| Race Code 5[d] | Hispanic/Latino | 1.14 | (0.61, 2.13) | 0.689 |
| I-SNP | Enrolled for at least one month in the institutional special needs plan | 0.71 | (0.64, 0.77) | <0.001 |
| HCC009 | Lung and other severe cancers | 0.70 | (0.55, 0.89) | 0.003 |

| | | | | |
|---|---|---|---|---|
| HCC027 | End-stage liver disease | 1.68 | (1.12, 2.51) | 0.011 |
| HCC028 | Cirrhosis of liver | 0.73 | (0.50, 1.05) | 0.086 |
| HCC039 | Bone/joint/muscle infections/necrosis | 1.11 | (0.89, 1.37) | 0.352 |
| HCC046 | Severe hematological disorders | 0.75 | (0.49, 1.15) | 0.183 |
| HCC047 | Disorders of immunity | 0.76 | (0.58, 0.99) | 0.038 |
| HCC048 | Coagulation defects and other specified hematological disorders | 1.11 | (0.97, 1.28) | 0.134 |
| HCC054 | Substance use with psychotic complications | 1.75 | (1.07, 2.86) | 0.026 |
| HCC055 | Substance use disorder, moderate/severe or substance use with complications | 1.15 | (0.96, 1.38) | 0.131 |
| HCC057 | Schizophrenia | 1.27 | (1.07, 1.52) | 0.007 |
| HCC077 | Multiple sclerosis | 1.35 | (1.00, 1.84) | 0.053 |
| HCC084 | Cardio-respiratory failure and shock | 1.07 | (0.96, 1.20) | 0.227 |
| HCC104 | Monoplegia, other paralytic syndromes | 0.34 | (0.16, 0.70) | 0.003 |
| HCC112 | Fibrosis of lung and other chronic lung disorders | 0.60 | (0.40, 0.91) | 0.017 |
| HCC114 | Aspiration and specified bacterial pneumonias | 1.02 | (0.86, 1.21) | 0.834 |
| HCC135 | Acute renal failure | 1.03 | (0.92, 1.16) | 0.566 |
| HCC158 | Pressure ulcer of skin with full thickness skin loss | 1.12 | (0.92, 1.35) | 0.262 |
| HCC167 | Major head injury | 1.55 | (1.10, 2.19) | 0.013 |
| HCC170 | Hip fracture/dislocation | 1.12 | (0.91, 1.37) | 0.280 |

[a]0-25% as baseline; [b]minimal or minor as baseline; [c]female as baseline; [d]unknown race as baseline
Significance was set at $p<0.05$. PIP: pharmacogenomic interaction probability; DDI: drug-drug interaction; COPD: chronic obstructive pulmonary disease; RAF: risk adjustment factor; I-SNP: institutional special needs plan.

**eTable 13. Expected length-of-stay ratios in the model for patients with diabetes**

| Variable | Description | Ratio | 95% C.I. | *P*-value |
|---|---|---|---|---|
| (Intercept) | Intercept | 8.45 | (6.60, 10.80) | <0.001 |
| Moderate PIP[a] | 26% to 50% | 1.05 | (0.99, 1.12) | 0.119 |
| High PIP[a] | >50% | 1.15 | (1.05, 1.25) | 0.002 |
| DDI Category 1[b] | Moderate, major, or contraindicated | 1.07 | (1.01, 1.13) | 0.031 |
| Hypertension | Hypertension flag | 0.98 | (0.91, 1.04) | 0.440 |
| Hyperlipidemia | Hyperlipidemia flag | 0.98 | (0.92, 1.04) | 0.544 |
| COPD | COPD flag | 0.98 | (0.92, 1.06) | 0.649 |
| Age | Age of a patient (2019) | 1.004 | (1.001, 1.007) | 0.008 |
| Income | Median household income per ZCTA level (standardized) | 1.08 | (1.05, 1.11) | <0.001 |
| RAF Score | Risk adjustment factor score | 1.03 | (1.00, 1.05) | 0.033 |
| Gender[c] | Gender of a patient | 1.08 | (1.01, 1.14) | 0.016 |
| D-SNP | Enrolled for at least one month in the dual-eligible special needs plan | 0.91 | (0.81, 1.03) | 0.144 |
| I-SNP | Enrolled for at least one month in the institutional special needs plan | 0.72 | (0.67, 0.77) | <0.001 |
| HCC009 | Lung and other severe cancers | 0.70 | (0.55, 0.88) | 0.003 |
| HCC010 | Lymphoma and other cancers | 1.25 | (0.99, 1.57) | 0.056 |
| HCC017 | Diabetes with acute complications | 0.85 | (0.72, 1.02) | 0.075 |
| HCC023 | Other significant endocrine and metabolic disorders | 0.94 | (0.84, 1.05) | 0.266 |
| HCC027 | End-stage liver disease | 1.32 | (0.94, 1.83) | 0.106 |

| | | | | |
|---|---|---|---|---|
| HCC033 | Intestinal obstruction/perforation | 1.07 | (0.91, 1.26) | 0.436 |
| HCC035 | Inflammatory bowel disease | 1.20 | (0.91, 1.58) | 0.191 |
| HCC039 | Bone/joint/muscle infections/necrosis | 1.10 | (0.94, 1.29) | 0.217 |
| HCC046 | Severe hematological disorders | 0.75 | (0.55, 1.03) | 0.071 |
| HCC048 | Coagulation defects and other specified hematological disorders | 1.07 | (0.96, 1.20) | 0.214 |
| HCC055 | Substance use disorder, moderate/severe or substance use with complications | 1.18 | (0.99, 1.42) | 0.072 |
| HCC072 | Spinal cord disorders/injuries | 1.25 | (0.93, 1.68) | 0.141 |
| HCC077 | Multiple sclerosis | 1.33 | (1.04, 1.69) | 0.022 |
| HCC082 | Respirator dependence/ tracheostomy status | 1.32 | (0.97, 1.79) | 0.078 |
| HCC099 | Cerebral hemorrhage | 1.24 | (0.98, 1.57) | 0.073 |
| HCC103 | Hemiplegia/hemiparesis | 1.07 | (0.96, 1.19) | 0.211 |
| HCC107 | Vascular disease with complications | 1.10 | (0.95, 1.26) | 0.208 |
| HCC134 | Dialysis status | 0.87 | (0.75, 1.02) | 0.081 |
| HCC135 | Acute renal failure | 1.02 | (0.93, 1.11) | 0.733 |
| HCC161 | Chronic ulcer of skin, except pressure | 1.11 | (0.99, 1.24) | 0.077 |

[a]0-25% PIP as baseline; [b]minimal or minor DDI as baseline; [c]female as baseline.
Significance was set at $P$<0.05. PIP: pharmacogenomic interaction probability; DDI: drug-drug interaction; COPD: chronic obstructive pulmonary disease; RAF: risk adjustment factor; I-SNP: institutional special needs plan.

**eTable 14. Expected length-of-stay ratios in the model for patients with hyperlipidemia**

| Variable | Description | Ratio | 95% C.I. | *P*-value |
|---|---|---|---|---|
| (Intercept) | Intercept | 6.46 | (5.06, 8.24) | <0.001 |
| Moderate PIP[a] | 26% to 50% | 1.08 | (1.02, 1.15) | 0.013 |
| High PIP[a] | >50% | 1.12 | (1.02, 1.22) | 0.014 |
| DDI Category 1[b] | Moderate, major, or contraindicated | 1.05 | (0.99, 1.11) | 0.096 |
| Hypertension | Hypertension flag | 1.02 | (0.96, 1.08) | 0.585 |
| Diabetes | Diabetes flag | 1.00 | (0.94, 1.06) | 0.974 |
| COPD | COPD flag | 1.05 | (0.98, 1.12) | 0.209 |
| Age | Age of a patient (2019) | 1.007 | (1.004, 1.010) | <0.001 |
| Income | Median household income per ZCTA level (standardized) | 1.07 | (1.04, 1.10) | <0.001 |
| RAF Score | Risk adjustment factor score | 1.03 | (1.00, 1.05) | 0.027 |
| Gender[c] | Gender of a patient | 1.06 | (1.00, 1.12) | 0.068 |
| I-SNP | Enrolled for at least one month in the institutional special needs plan | 0.75 | (0.71, 0.80) | <0.001 |
| HCC009 | Lung and other severe cancers | 0.74 | (0.58, 0.94) | 0.015 |
| HCC023 | Other significant endocrine and metabolic disorders | 0.87 | (0.77, 0.97) | 0.017 |
| HCC028 | Cirrhosis of liver | 0.72 | (0.51, 1.01) | 0.059 |
| HCC046 | Severe hematological disorders | 0.62 | (0.45, 0.86) | 0.004 |
| HCC055 | Substance use disorder, moderate/severe or substance use with complications | 1.18 | (1.01, 1.39) | 0.038 |
| HCC058 | Reactive and unspecified psychosis | 1.07 | (0.98, 1.15) | 0.123 |

| | | | | |
|---|---|---|---|---|
| HCC072 | Spinal cord disorders/injuries | 1.15 | (0.88, 1.51) | 0.307 |
| HCC077 | Multiple sclerosis | 1.33 | (1.04, 1.69) | 0.022 |
| HCC082 | Respirator dependence/ tracheostomy status | 1.51 | (1.11, 2.07) | 0.009 |
| HCC084 | Cardio-respiratory failure and shock | 1.07 | (0.97, 1.19) | 0.184 |
| HCC099 | Cerebral hemorrhage | 1.15 | (0.89, 1.48) | 0.284 |
| HCC104 | Monoplegia, other paralytic syndromes | 0.58 | (0.36, 0.94) | 0.027 |
| HCC112 | Fibrosis of lung and other chronic lung disorders | 0.78 | (0.56, 1.07) | 0.125 |
| HCC167 | Major head injury | 1.06 | (0.84, 1.35) | 0.618 |
| HCC170 | Hip fracture/dislocation | 1.22 | (1.03, 1.45) | 0.022 |

[a]0-25% PIP as baseline; [b]minimal or minor DDI as baseline; [c]female as baseline.
Significance was set at $P<0.05$. PIP: pharmacogenomic interaction probability; DDI: drug-drug interaction; COPD: chronic obstructive pulmonary disease; RAF: risk adjustment factor; I-SNP: institutional special needs plan.

### eTable 15. Expected length-of-stay ratios in the model for patients with hypertension

| Variable | Description | Ratio | 95% C.I. | *P*-value |
|---|---|---|---|---|
| (Intercept) | Intercept | 12.45 | (11.24, 13.79) | <0.001 |
| Moderate PIP[a] | 26% to 50% | 1.03 | (0.95, 1.12) | 0.478 |
| High PIP[a] | >50% | 1.22 | (1.10, 1.35) | <0.001 |
| DDI Category 1[b] | Moderate, major, or contraindicated | 1.02 | (0.95, 1.10) | 0.522 |
| Hyperlipidemia | Hyperlipidemia flag | 1.02 | (0.95, 1.10) | 0.617 |
| Diabetes | Diabetes flag | 0.93 | (0.86, 1.00) | 0.058 |
| COPD | COPD flag | 1.04 | (0.95, 1.13) | 0.403 |
| Income | Median household income per ZCTA level (standardized) | 1.08 | (1.04, 1.11) | <0.001 |
| RAF Score | Risk adjustment factor score | 1.01 | (0.99, 1.04) | 0.321 |
| I-SNP | Enrolled for at least one month in the institutional special needs plan | 0.73 | (0.68, 0.79) | <0.001 |
| HCC039 | Bone/joint/muscle infections/necrosis | 1.17 | (0.98, 1.40) | 0.091 |
| HCC084 | Cardio-respiratory failure and shock | 1.08 | (0.97, 1.21) | 0.148 |
| HCC135 | Acute renal failure | 1.07 | (0.97, 1.17) | 0.175 |

[a]0-25% PIP as baseline; [b]minimal or minor DDI as baseline.
Significance was set at $P<0.05$. PIP: pharmacogenomic interaction probability; DDI: drug-drug interaction; COPD: chronic obstructive pulmonary disease; RAF: risk adjustment factor; I-SNP: institutional special needs plan.
